# How do the public interpret COVID-19 swab test results? Comparing the impact of official information about results and reliability used in the UK, US and New Zealand: a randomised, controlled trial

**DOI:** 10.1101/2020.12.04.20243840

**Authors:** Gabriel Recchia, Claudia R. Schneider, Alexandra L.J. Freeman

**Author notes:** Corresponding Author Alexandra Freeman. These authors contributed equally to the work (Gabriel Recchia, Claudia R. Schneider). Contributor statement GR conceived of the experiment; GR, CRS and ALJF designed the questionnaire; GR & CRS carried out the quantitative analysis; ALJF analysed the free text; all authors wrote the manuscript.

## Abstract

**Objectives:** To assess the effects of different official information on public interpretation of a personal COVID-19 PCR (‘swab’) test result.

**Design:** A 5x×2 factorial, randomised, between-subjects experiment, comparing four wordings of information about the test result and a control arm of no additional information; for both positive and negative test results.

**Setting:** Online experiment using recruitment platform Respondi.

**Participants:** UK participants (n=1,744, after a pilot of n=1,657) collected by quota sampling to be proportional to the UK national population on age and sex.

**Interventions:** Participants were given a hypothetical COVID-19 swab test result for ‘John’ who was presented as having a 50% chance of having COVID-19 based on symptoms alone. Participants were randomised to receive either a positive or negative result for ‘John’, then randomised again to receive either no more information, or text information on the interpretation of COVID-19 test results copied from the public websites of the UK’s National Health Service, the US’s Centers for Disease Control, New Zealand’s Ministry of Health, or a modified version of the UK’s wording incorporating uncertainty. Information identifying the source of the wording was removed.

**Main outcome measures:** Participants were asked “What is your best guess as to the percent chance that John actually had COVID-19 at the time of his test, given his result?”; questions about their feelings of trustworthiness in the result, their perceptions of the quality of the underlying evidence, and what action they felt ‘John’ should take in the light of his result.

**Results:** Of those presented with a positive COVID-19 test result for ‘John’, the mean estimate of the probability that he had the virus was 73%; for those presented with a negative result, 38%. There was no main effect of information (wording) on these means. However, those participants given the official information on the UK website, which did not mention any uncertainty around the test result, were significantly more likely to give a categorical (100% or 0%) answer (for positive result, *p* <.001; negative, *p* =.006). When asked how much they agreed that ‘John’ should self-isolate, those who were told his test was positive agreed to a greater extent (mean 86 on a 0-100 scale), but many of those who were told he had a negative result still agreed (mean 51). There was also an interaction between wording and test result (*p* < 0.001), with those seeing the New Zealand wording about the uncertainties of the test result significantly more likely to agree that he should continue to self-isolate after a negative test than those who saw the UK wording (*p* =.01), the experimental wording (*p* =.02) or no wording at all (*p* =.003). Participants rated positive test results more trustworthy and higher quality of evidence than negative results.

**Conclusions:** The UK public perceive positive test results for COVID-19 as more reliable and trustworthy than negative results without being given any information about the reliability of the tests. When additionally given the UK’s current official wording about the interpretation of the test results, people became more likely to interpret the results as definitive. The public’s assessment of the face value of both the positive and negative test results was generally conservative. The proportion of participants who felt that a symptomatic individual who tests negative definitely should *not* self-isolate was highest among those reading the UK wording (17.4%) and lowest among those reading the New Zealand wording (3.8%) and US wording (5.1%).

**Pre-registration and data repository:** pre-registration of pilot at osf.io/8n62f, pre-registration of main experiment at osf.io/7rcj4, data and code in https://osf.io/pvhba/.

**What is already known on this topic:**

- Different countries have had different approaches to conveying the meaning of a COVID-19 swab test result, particularly regarding the uncertainties inherent in the result due to limitations of specificity and sensitivity.
- Previous research has suggested that people’s trust and understanding is not affected by conveying quantified uncertainties numerically, but that perceptions of the quality of the underlying evidence can affect trust.
- It is not known whether the different wordings around COVID-19 test uncertainties are likely to affect people’s trust in, or behavioural response to, the results they receive.

**What this study adds:**

- This study provides the first empirical evidence to our knowledge of the responses the public have to COVID-19 swab test results.
- It suggests that the public have a higher degree of trust and confidence in positive swab test results than negative when they are not given any other information accompanying the result. The experimental wording that we created for this study appeared to boost their trust in and assessment of quality of positive test results, but did not change their lower ratings of negative results.
- The wording used by the UK’s National Health Service, which does not include any cues of uncertainty in the result, was more likely to lead people to definitive (100% or 0%) answers to questions about the meaning of the result.
- The wording used by New Zealand’s Ministry of Health, which is more explicit about the reliability of the tests, appears to lead people to be more cautious about recommending that a test participant with a negative test (but still symptomatic) no longer needs to self-isolate.

## INTRODUCTION

PCR or ‘swab’ tests for COVID-19 give policymakers vital information about the prevalence of the virus, and also help individuals take appropriate action such as self-quarantining and initiating contact-tracing. However, the sensitivity and specificity of the tests (especially in ‘real world use’ scenarios) are not 100% and hence people taking action on the basis of the information need to be aware of the potential that for an individual, a single negative test may not be an ‘all clear’. Watson et al. (1) created a tool to help with the interpretation of such tests, given known sensitivity, specificity and prevalence values, suggesting that a positive test result with a prior suspicion of COVID of 50% should be interpreted as around a 93% chance that the patient had COVID-19 at the time they were tested, and a negative test as around a 24% chance that the patient had COVID-19 (1). A recent assessment of the UK Office for National Statistics estimated a false positive rate of under 0.005% (2), suggesting that even more confidence about the interpretation of a positive test result may be warranted.

However, the wording provided to support public interpretations of the test results varies from country to country.

In the UK, the National Health Service website on test interpretation expresses no uncertainty (e.g. “*A positive result means you had coronavirus when the test was done*” and “*A negative result means the test did not find coronavirus*”)(3).

On an equivalent US website from the Centers for Disease Control, the phrasing is very slightly less certain (e.g. “*If you test positive, know what protective steps to take to prevent others from getting sick*” and “*If you test negative, you probably were not infected at the time your sample was collected. The test result only means that you did not have COVID-19 at the time of testing*”)(4,5).

By contrast, in New Zealand, a much longer wording is provided online from the Ministry of Health around test interpretation which includes far more information about the uncertainties inherent in the test (e.g. “*A recent laboratory study found that different COVID-19 testing kits correctly detected COVID-19 in samples more than 95% (and frequently 100%) of the time. When tests were done on samples without the virus, the tests correctly gave a negative result 96% of the time. But it is important to remember that tests don’t work as well in the real world*.*”* and *“The viral test for COVID-19 is much better at correctly identifying people who don’t have COVID-19 (this is known as a higher ‘specificity’). We expect very few (if any) false positive test results (a false positive being a positive test result for someone who does not have the disease)*.”)(6)

Concern has been expressed that information provided to patients without uncertainty may cause unwarranted confidence in the results, particularly negative results(7). This study, therefore, set out to test the effects of these different approaches to communicating COVID-19 swab test results on a public audience.

After a pilot study (see Supplementary Information) we created an experimental version of the information, based on the UK’s NHS website wording but containing elements to make the uncertainties explicit, based loosely on that provided on the New Zealand Ministry of Health website.

## METHODS

The study was conducted with ethical oversight from the University of Cambridge Psychology Research Ethics Committee (PRE.2020.034 with amendment 15^th^ September 2020). Participants aged 18+ were recruited for the main study online through the ISO-accredited company Respondi and directed to a questionnaire in Qualtrics (see Supplementary Information for details on the pilot). All participants gave informed consent before participation and were randomised using the ‘randomise’ function in Qualtrics.

Characteristics of the participants in the main study are shown in Table 1. The questionnaire took a median of 14 minutes for participants to complete, and they were paid £0.75 for their participation. Analysis was conducted in R (version 3.6).

**Table 1:**
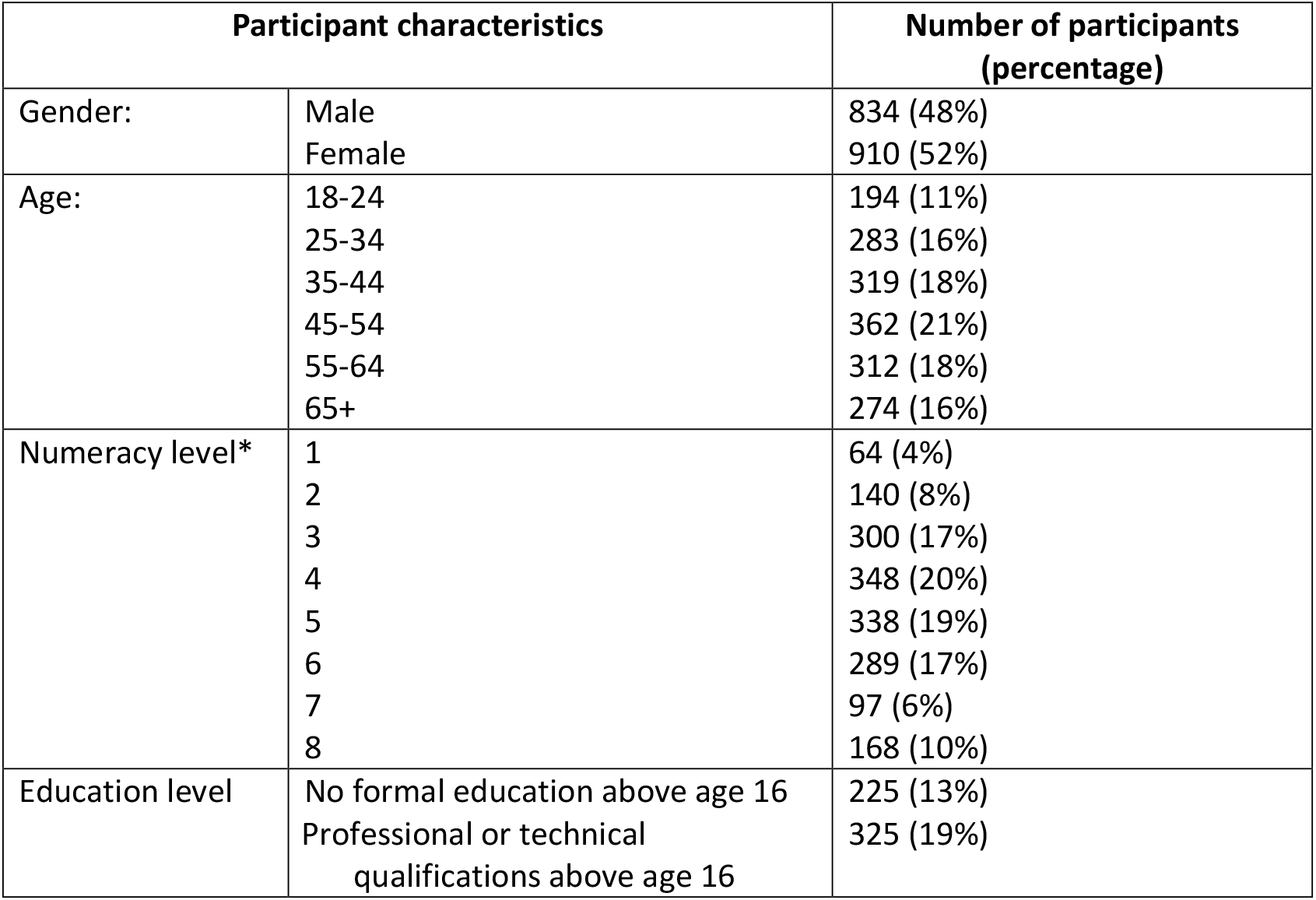
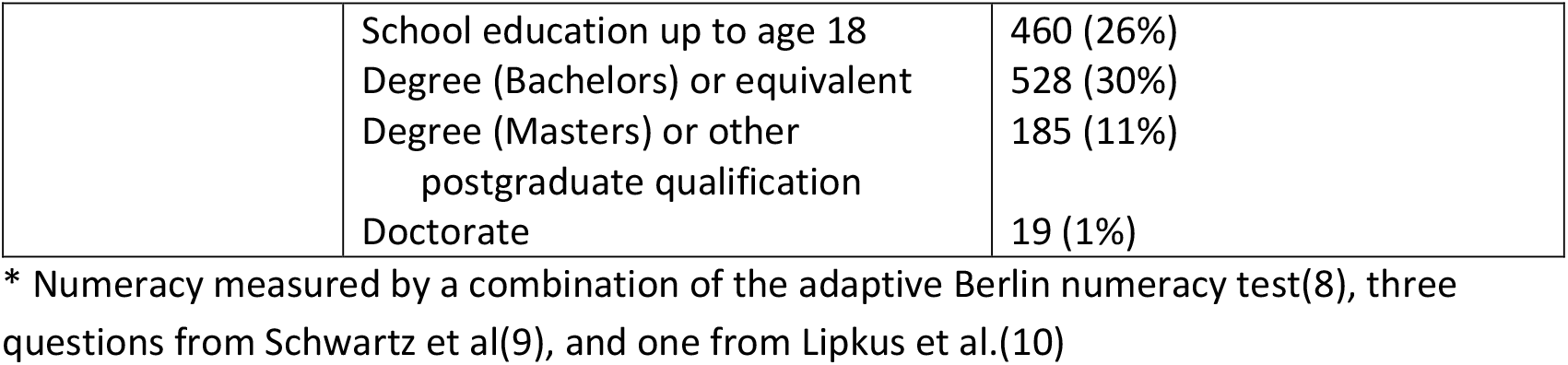
Characteristics of the participants

Participants were presented with the scenario “John has been feeling ill. Based on his symptoms alone, a knowledgeable doctor believes that John has a 50-50 chance of having COVID-19. John takes the COVID-19 (swab) test to see whether he currently has the virus, and it is”… with the final word of the sentence being *positive* or *negative*, depending on whether they had been randomized into the *positive* or *negative* ‘test result’ condition. Unlike in the pilot, we also placed the final word of the sentence in bold font, to increase the likelihood that participants would read and remember the result. Participants were then further randomised to one of five ‘information’ conditions. Those in a control condition received no further information; others were given the explanatory text around interpreting COVID-19 test results used on official websites produced by one of the following: the UK’s National Health Service, the US’s Centers for Disease Control, New Zealand’s Ministry of Health (with any identifying information of where the text had come from removed), or an experimental wording which added information about the possibility of a false positive or false negative result to the original UK wording and attempted to make the guidelines clearer about when a person with a negative test result should still self-isolate. The full texts given to participants are shown in Supplementary Information.

Participants were then asked a series of questions, including “What is your best guess as to the percent chance that John actually had COVID-19 at the time of his test, given his result?” and questions about: their impression of the accuracy, reliability, certainty and trustworthiness of the result (combined into a single index, ‘trustworthiness’); the quality of the evidence it was based on; how much they agreed with a statement that ‘John’ should continue to self-isolate as a result of his test; their confidence in their estimate of the chance that ‘John’ had COVID-19; and how clear and easily understood the information was that they were given (the last questions about clarity and ease of understanding were not asked in the control condition). They were also asked a number of demographic questions and questions about their experience with, and perception of the risk of, COVID-19. After the question on whether they thought the test recipient should self-isolate, participants were asked why they answered the way they did and given a free text response box. Timers were also set to measure the amount of time participants spent reading the information and answering the sets of questions. Full details of the questions and their answer options are shown in the Supplementary Information.

In the pilot experiment, participants were asked to type in their best guess as to the percent chance that John actually had COVID-19 at the time of his test. For the main experiment this was changed to a slider and labels added to confirm ‘0%: John definitely **did not** have COVID-19’ and ‘100%: John definitely **did** have COVID-19’ because in the pilot, close to half of participants typed in ‘50%’ as their answer, which is well known as a response to people not feeling comfortable working on a numeric scale that can be mitigated by providing a slider (11,12).

### Power calculation

Based on the effect sizes achieved in the pilot, we calculated the number of participants required to achieve 90% power to detect an effect size of *η* _*p*_^*2*^ =.009 (f =.097), and adjusted our total required sample upwards on the assumption that the same proportion of participants would fail the attention check as did in the pilot. Consequently, we sampled 2,041 participants. Of these, 297 did not pass the attention check and were excluded from all analyses. Final analytical samples in each condition were: Control positive: 156, negative: 200; US positive: 165 negative: 177; New Zealand positive: 186, negative: 159; UK positive: 165, negative: 178; Experimental positive: 200, negative: 158. We pre-registered 2 main hypotheses and 2 additional weaker secondary hypotheses (for details see osf.io/7rcj4) based on the findings of the pilot.

### Pre-registered hypotheses

In the pilot, we found that for participants in the ‘positive’ test result condition, their estimates of the probability that John actually had COVID-19 were lower in those shown the UK or US information than in those shown the New Zealand information or no information at all. We found this surprising, but thought this might be because the UK and US information did not explicitly note that positive test results were more reliable than negative test results, potentially causing participants’ pre-existing scepticism about negative test results (or test results in general) to affect their views of positive test results as well. Therefore, our first primary hypothesis (H1) was that messaging which does not specifically contrast the low level of uncertainty associated with a positive test result with the relatively high level of uncertainty associated with a negative test result will lead participants to estimate that the true probability of having COVID-19 after a positive test result is lower compared with participants who saw other messaging.

Specifically, we pre-registered that we would expect an interaction between message type and test result (positive vs. negative), such that when estimating the true probability of having COVID-19 after a positive test result, participants who view messages that do not emphasize differences in certainty levels (the wording from the UK and US) will estimate that the true probability of having COVID-19 is lower as compared with participants in: (1) a control condition in which no message is shown at all, (2) the condition with the messaging from New Zealand, and (3) our ‘experimental’ information condition which did emphasize differences in the interpretation of positive and negative test results.

Our second primary hypothesis (H2) was that participants who are shown no messaging at all, and participants in the experimental information condition, would similarly perceive positive test results to have higher quality of evidence than negative test results (in contrast to the those seeing the UK and US information, where we did not expect to find this result). This hypothesis was likewise based on the findings of the pilot study, specifically, the analysis of responses to the question ‘How high or low do you think the quality of the evidence behind John’s test result is?’, which we used as the quality-of-evidence measure in the present study.

Specifically, we pre-registered that we expected an interaction between message wording and test result, such that for participants presented with positive test results in the control and ‘experimental’ information conditions, positive test results would be rated as having higher quality of evidence than negative test results, while we did not expect to find evidence of this for participants in conditions that do not emphasize differences in certainty levels (the UK and US).

Finally, we pre-registered two secondary hypotheses. H3 was as in H2, but with respect to the trustworthiness of the test result rather than the quality of evidence behind the test. (Although post-hoc analysis of the pilot data did suggest a difference in trustworthiness between positive and negative test results for those viewing the US message, we theorized that this specific difference might not replicate, given its inconsistency with the patterns of results that led us to H1 and H2.) H4 was as in H2, but with respect to participants’ perception that John should isolate himself from other people, which was not investigated in the pilot.

## RESULTS

5 (message wording) x 2 (test result) ANOVAs were conducted for each primary dependent variable: participants’ estimates of John’s chance of having COVID, the quality of evidence behind John’s test result, the trustworthiness of John’s test result, and level of agreement with the statement “John should now isolate himself from other people”. Aligned ranks transformation ANOVAs were used for ‘chance of COVID’ estimates and levels of agreement due to deviations from normality. As aligned rank transformation ANOVAs must always include interaction terms, these models always included the wording * test result interaction term. For other ANOVAs, we ran the model with the interaction term (using type 3 sums-of-squares), but when there was no interaction, we excluded the interaction term and reported a main-effects-only model (type 2 sums-of-squares).

### How likely did participants think it was that the test recipient actually had COVID-19 given a positive or negative test result?

Overall 25.4% of participants gave an answer between 48-52%, a phenomenon that is commonly observed and is attributed to ‘50-50’ being akin to ‘I don’t know’(11). There was no difference in the prevalence of this answer between the different wording groups with the exception of participants who saw the New Zealand wording, of whom only 18.5% gave answers between 48% and 52%.

An aligned ranks transformation ANOVA found a main effect of test result (*F*(1,1734) = 976.2, *p* <.001, *η*^*2*^ = 0.99. The mean estimate of the likelihood of the test recipient having COVID-19 given that he received a positive result was 73.0% (*SD =* 22.6%, median = 75%), and the mean estimate for him having the virus given a negative result was 38.4% (*SD =* 25.1%, median = 50%). However, there was no main effect of message wording and no interaction. Exploratory one-way ANOVAs likewise found no main effect of message wording in either those who were presented with a positive test result or those presented with a negative result. See Figure 1.

**Figure 1:**
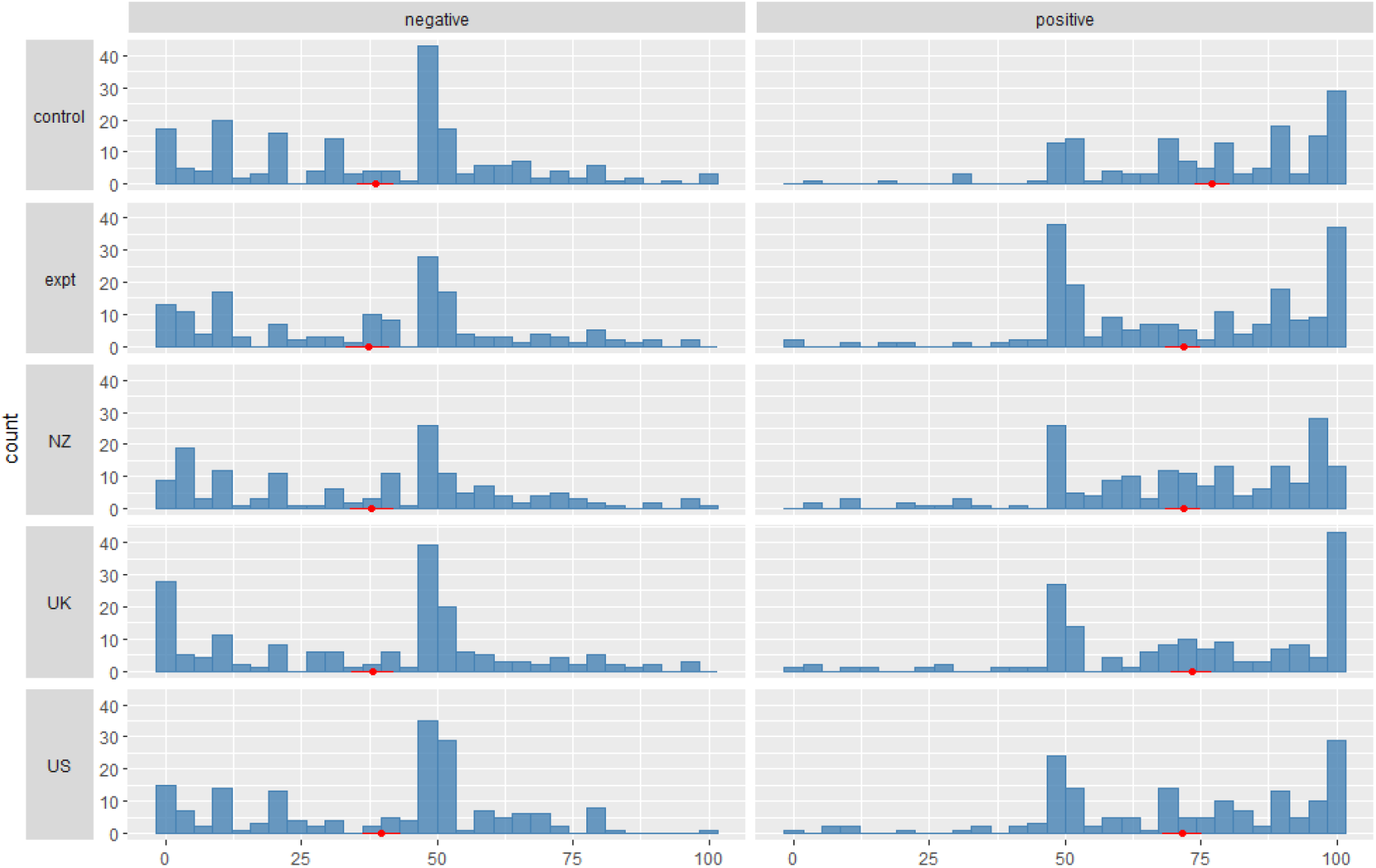
Participants’ estimates of the chances of the test recipient John actually having COVID-19, given either a negative (left hand side) or positive (right hand side) test result, and with either no accompanying explanatory text (control), official text from one of three different countries (New Zealand, UK, US), or an experimental text (expt) based on the UK wording but with added clarification about the test uncertainties. The large hump around 50% is often interpreted as a response from participants who ‘don’t know(11)’. Red markers indicate mean and 95 percent confidence interval.

However, further exploratory analysis revealed that the distributions behind the means of each wording group were different. Participants seeing the UK wording were more likely to give a categorical ‘100%’ or ‘0%’ answer in the positive and negative test scenarios than were those who saw other wordings (for positive test results meaning 100%, *X*^2^(4, *N* = 872) = 22.1, *p* <.001; for negative test results meaning 0%, *X*^2^(4, *N* = 872) = 14.4, *p* =.006; for any test result meaning 0% or 100%, *X*^2^(4, *N* = 1744) = 32.0, *p* <.001). See Figure 2a&b. Follow-up *X*^2^ tests comparing the UK wording condition to each other group in turn found that those in the UK wording group were significantly more likely to give a categorical response than every other group, and those in the NZ wording group were significantly less likely to give a categorical response than any other group, all tests *p* <.05.

**Figure 2a&b.**
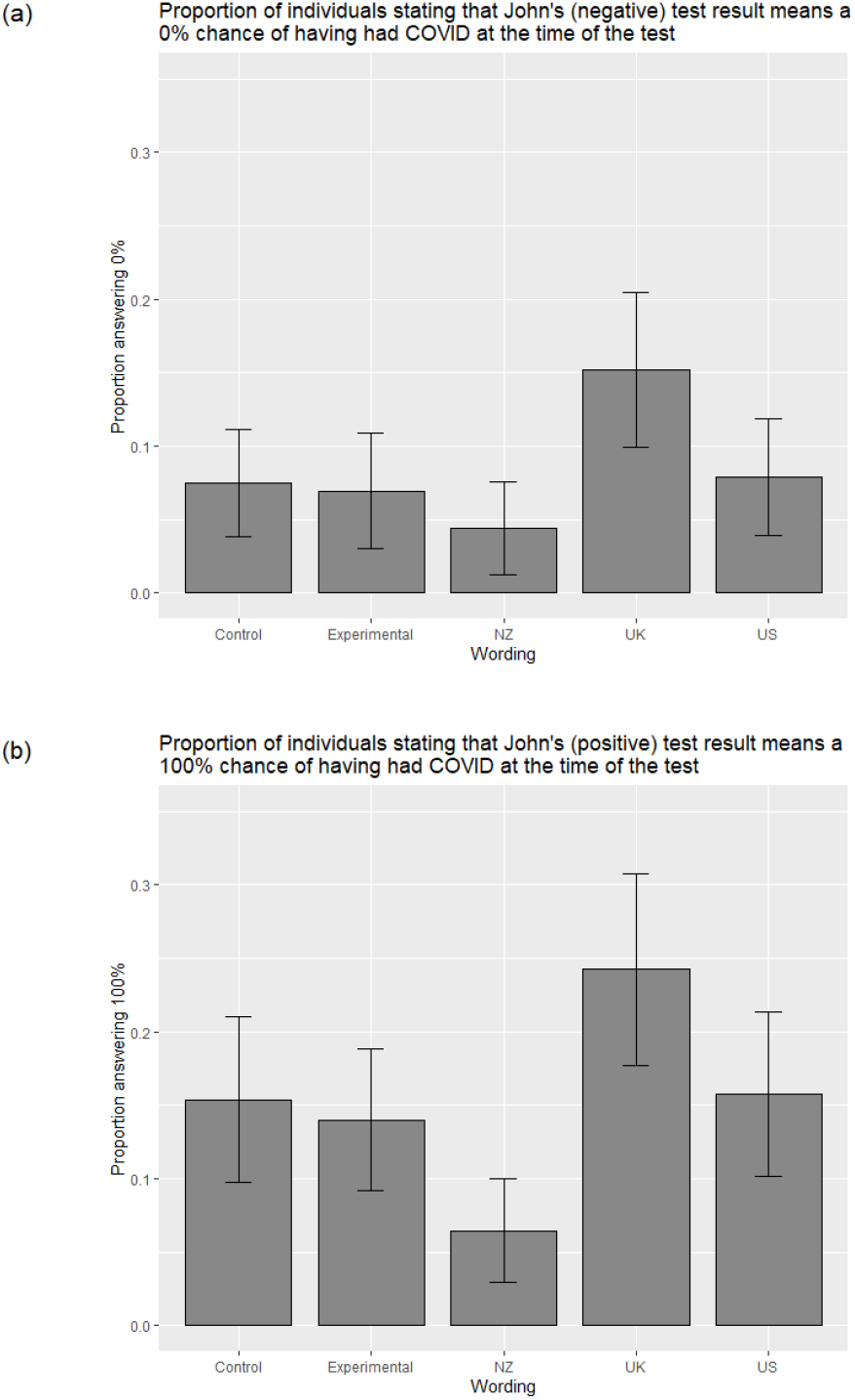
The proportion of participants giving categorical 100% (a) or 0% (b) responses as to what John’s viral test result means in each message wording group, for those who were told that John’s result was (a) positive and (b) negative

Because the mean and median answers may be skewed by the people answering ‘50%’ as a signal of not knowing, rather than as an actual estimate, and because estimates of the true positive and negative predictive values of COVID-19 tests are uncertain(1), it is difficult to compare these average answers with the estimated actual chance of having COVID-19.

After removing participants who answered between 48-52% and also gave a rating of their confidence in their answer at 50% or below – which possibly indicates that they were indicating ‘I don’t know’ rather than that they thought the likelihood of the recipient having COVID-19 was around 50% – the average percentages were 74.3% for a positive test result and 37.1% for a negative test result. A factorial ANOVA on these participants did not reveal a main effect of wording nor an interaction with test result. Further analysis using two one-way ANOVAs for positive and negative test result respectively confirmed this pattern, i.e. no significant differences between the wording conditions emerged.

The results therefore did not confirm hypothesis H1, based on our pilot, that estimates of the positive predictive value (the likelihood that someone testing positive actually has COVID-19) would be lower in the UK and US wording conditions than in the others.

However, they do show that those seeing the UK wording took both the positive and negative results as more definitive than those in the other wording conditions (including the control group, which yields insight into people’s beliefs about test result interpretation prior to being shown further information).

### How much confidence did participants have in their answer regarding the likelihood that he actually had COVID-19?

An exploratory ANOVA found a main effect of test result, *F*(1, 1738) = 48.6, *p* <.001, *η*^*2*^ = 0.027, with greater confidence for positive test results (*M* = 72.3, *SD* = 24.7) than negative (*M* = 64.0, *SD* = 26.8). Likewise, there was a main effect of wording, *F*(4, 1738) = 4.1, *p* =.003, *η*^*2*^ = 0.009; post-hoc Tukey tests revealed that participants seeing the New Zealand wording were significantly less confident in their estimates of the likelihood that the test recipient had COVID-19 (*M* = 64.0, *SD* = 26.2) than those viewing the UK wording (*M* = 70.3, *SD* = 26.4), *p* =.004, or the control (*M* = 69.9, *SD* = 25.0), *p* =.004. See Figure 3.

**Figure 3:**
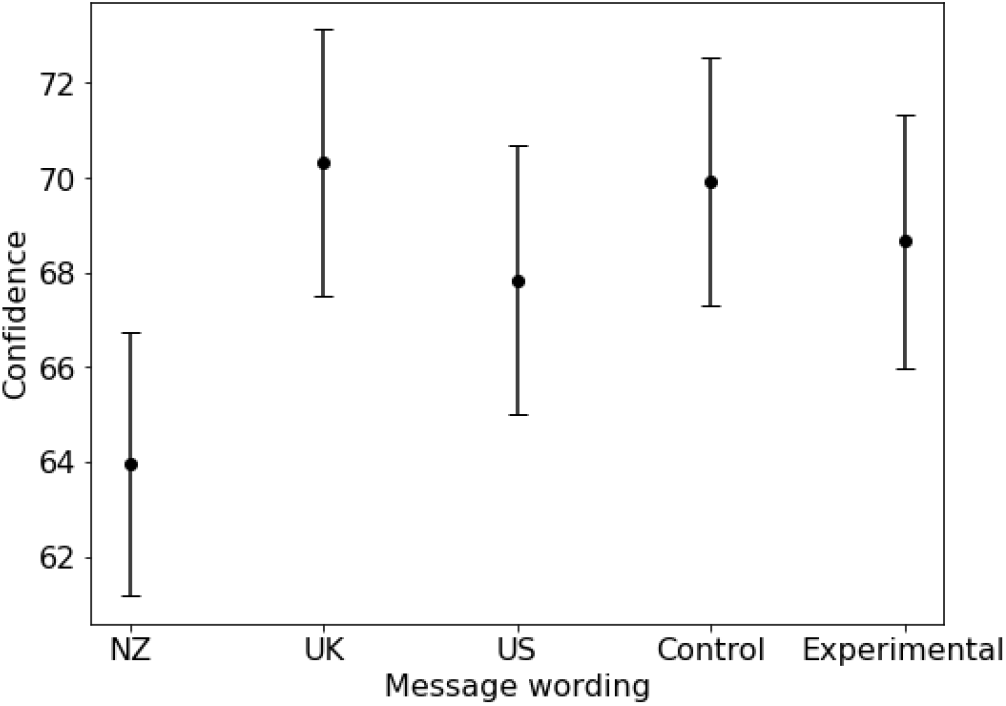
Participant confidence in their own estimates of the likelihood that the test recipient had COVID-19, by wording group.

### How did participants think the test recipient should behave after receiving either the positive or negative result?

Participants were asked “Given John’s test result, how much do you agree or disagree with the statement that ‘John should now isolate himself from other people’?”. They answered on a slider scale between ‘completely disagree’ to ‘completely agree’, which was converted to 0-100 for analysis. See Figure 4.

**Figure 4:**
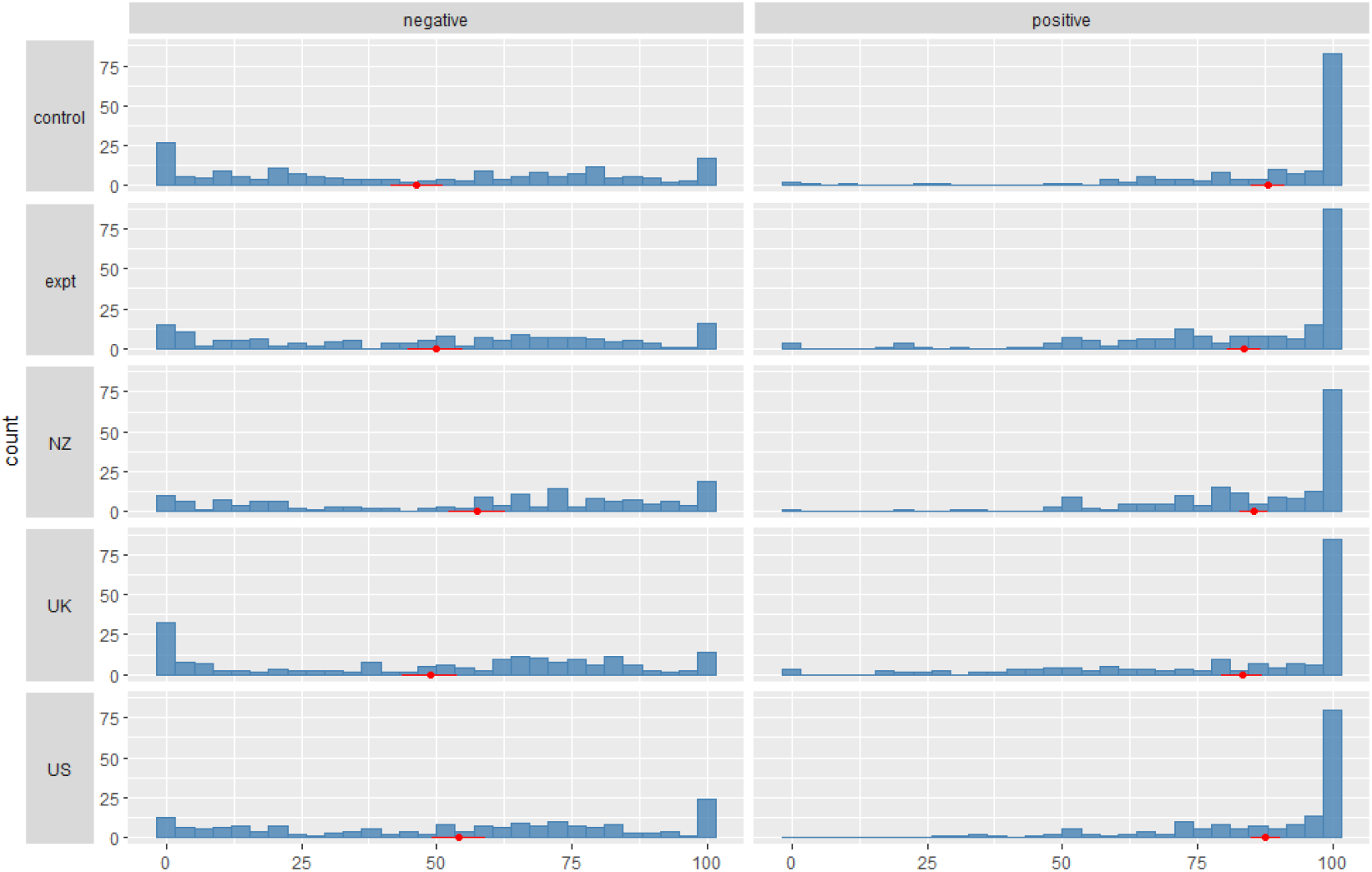
Participants’ amount of disagreement (0) or agreement (100), with the statement that the test recipient John should continue to self-isolate given either a negative (left hand side) or positive (right hand side) test result, and with either no accompanying explanatory text (control), official text from one of three different countries (New Zealand, UK, US), or an experimental text (expt) based on the UK wording but with added clarification about the test uncertainties. Red markers indicate mean and 95 percent confidence interval.

Participants who were given a positive result for John on average very much agreed with the statement (*M* = 85.5, *SD =* 20.7, median = 97). Participants who saw the negative condition showed a more divergent opinion (*M =* 51.1, *SD =* 33.8, median = 58.5). Aligned ranks transformation ANOVA revealed was a main effect of test result (*F*(1,1734) = 699.3, *p* <.001, *η*^*2*^ = 0.883) and a main effect of wording (*F*(4,1734) = 8.27, *p* <.001, *η*^*2*^ = 0.058). Aggregated over test result, post-hoc Mann-Whitney tests suggested more agreement with the statement among participants viewing the New Zealand wording (median 81) than among those viewing the control (median 77, *p* =.03) or the UK wording (median 75, *p* =.02). There was also a significant interaction (*F*(4,1734) = 8.51, *p* <.001). For example, of those shown negative test results, post-hoc Mann Whitney tests suggested that those viewing the New Zealand wording were more likely to believe that John should still isolate (median 65) than those viewing the experimental (median 55.5, *p =*.02) or the UK (median 60, *p* =.01) wording, but these differences were not observed for those shown positive test results.

However, the means and medians again obscure other important differences between message wording groups. Participants were more likely to express maximal disagreement with the statement “John should now isolate himself from other people” after hearing that John has been experiencing symptoms but tested *negative*, if they were presented with the UK message (*X*^2^(4, *N* = 872) = 23.9, *p* <.001): 17.4% of those in the UK wording group expressed maximal disagreement, in contrast to 3.8% (NZ wording), 5.1% (US wording), 8.9% (experimental wording), and 11.0% (control). Compared head-to-head with the control group, only the New Zealand wording group was less likely to express maximal disagreement, *X*^2^(2, *N* = 359) = 5.5, *p* =.02. Head-to-head comparisons between the UK wording condition and each other group found that those in the UK wording group were significantly more likely to express maximal disagreement than every other group except for the control, for which there was no significant difference. Participants who heard that John had tested *positive* were no more likely to express maximal agreement in one wording condition over another (*X*^2^(4, *N* = 872) = 5.3, *p* =.26).

The results partially confirm hypothesis H4, that there would be an interaction between wording and test result. Those shown a positive test result were more likely to agree that the recipient should self-isolate, but the differences between the wording groups were not as hypothesised. Also, rather than finding a difference between control/experimental and US/UK we found a difference between New Zealand and experimental/UK for those shown the negative result, suggesting that the experimental wording was not as effective as the New Zealand wording at implying that people should be cautious with their behaviour (and self-isolate if they still had symptoms) despite a negative result.

Free text answers to why participants felt the test recipient should self-isolate were thematically coded by one of the authors into one of nine themes: mistakenly remembering the direction of the test result; scepticism about the seriousness or even existence of COVID-19; uncertainty about the validity of the test result (including quality of evidence concerns); John’s test result means he doesn’t need to self-isolate; concern for the mental health, personal freedom, practical or financial implications of self-isolation; scepticism about the effectiveness of self-isolation; John’s symptoms mean he should self-isolate; John’s test result means he should self-isolate; general comments about self-isolation being important for public health, or ‘better safe than sorry’; other/not sure/forgotten result. Where more than one reason was given, the response was scored on the first mentioned. See Figure 5.

**Figure 5:**
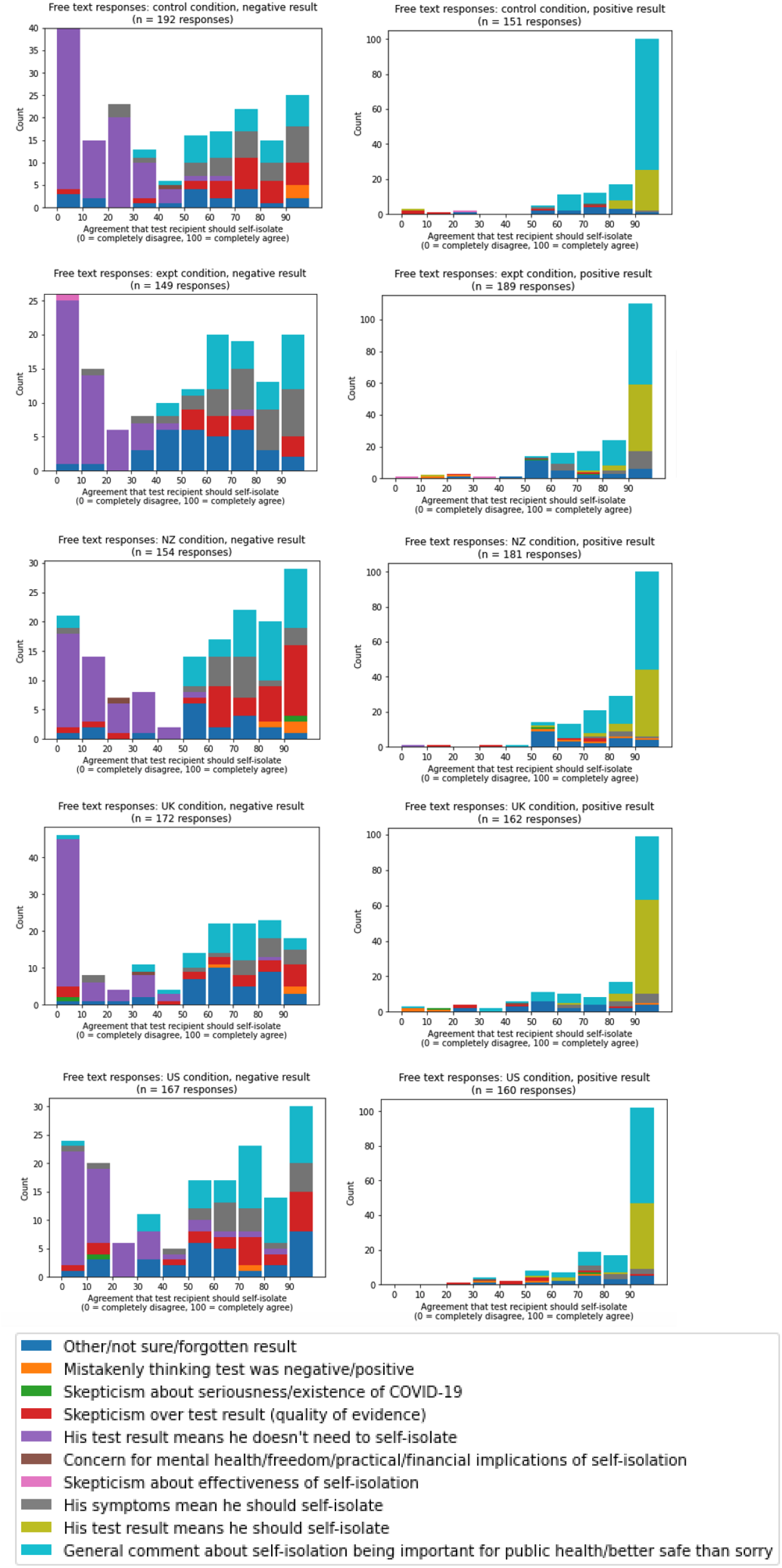
The reasons participants gave for their response to how much they agreed with the statement that the test recipient (who was symptomatic) should isolate after receiving his test result, split by the result they were given for the recipient (positive or negative) and the additional information that they received alongside the test result (control group received no additional information to help interpret the result).

### How trustworthy and high quality did participants think the test result was?

In both our measures of trustworthiness of the result and the quality of evidence of the result, we found that participants scored the positive test results (quality of evidence: *M* = 5.21, *SD* = 1.34; trustworthiness: *M* = 5.17, *SD* = 1.39) higher than negative test results (quality of evidence: *M* = 4.83, *SD* = 1.34; trustworthiness: *M* = 4.75, *SD* = 1.38), showing a main effect of test result for both quality of evidence (*F*(1,1738) = 37.09, *p* <.001, *η*^*2*^ = 0.021) and perceived trustworthiness (*F*(1,1738) = 40.42, *p* <.001, *η*^*2*^ = 0.023). See Figure 6a&b.

**Figure 6a&b:**
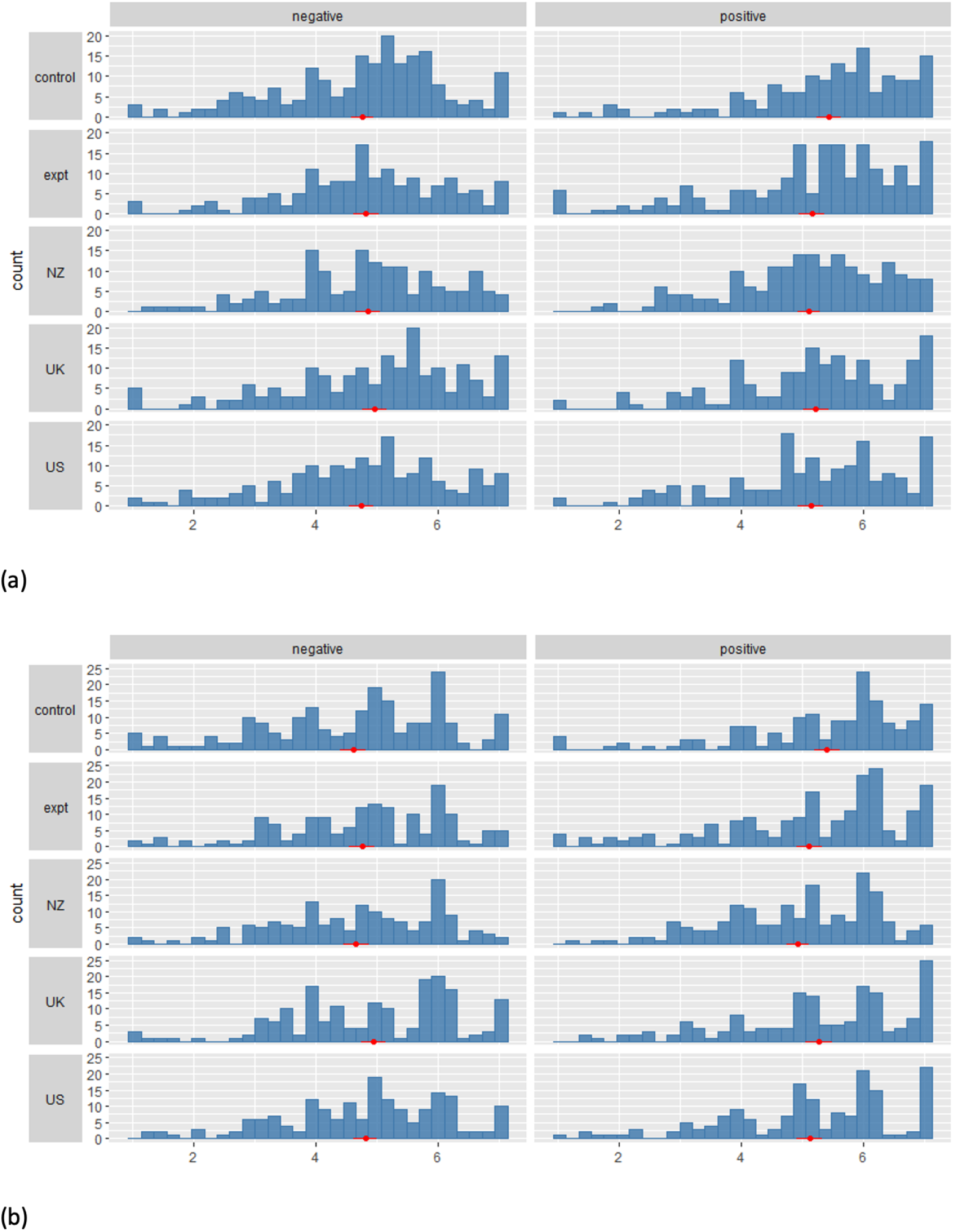
Participants’ scoring on a 1-7 Likert scale of the quality of the evidence (a) or the trustworthiness/certainty/accuracy/reliability (b) of the swab test given either a negative (left hand side) or positive (right hand side) test result, and with either no accompanying explanatory text (control), official text from one of three different countries (New Zealand, UK, US), or an experimental text (expt) based on the UK information but with added clarification about the test uncertainties. Red markers indicate mean and 95 percent confidence interval.

For perceived trustworthiness we additionally find a very small main effect of wording (*F*(4,1738) = 2.41, *p* =.048, *η*^*2*^ = 0.006), with participants shown the New Zealand wording (*M*=4.81, *SD*=1.28) rating it slightly less trustworthy than the UK wording (*M*=5.1, *SD*=1.39, p=.02). However, this effect is only significant when the interaction term is removed from the model, and was not found in the pilot. We did not find a main effect of wording for quality of evidence, and did not find significant interactions for either quality of evidence or perceived trustworthiness.

In pre-registered exploratory analyses, we found that higher numeracy was associated with higher quality of evidence perception (*b*=0.07, *SE*=0.02, p<.001, R^2^=0.01) and higher trustworthiness ratings (*b*=0.09, *SE*=0.02, p<.001, R^2^=0.01). If numeracy was added to the main effects models of test result and test wording, the main effect of test result stayed significant for both perceived trustworthiness as well as quality of evidence perception (both p<.001); likewise numeracy emerged as a significant predictor in those models (both p<.001) for both quality and trust perception.

We also found a positive correlation between people’s agreement that ‘John’ should self-isolate after a positive test and quality of evidence perception (r=0.51, *t*=17.68, df=870, p<.001) as well as perceived trustworthiness (r=0.53, *t*=18.63, df=870, p<.001). With a negative test, we found a negative correlation between people’s agreement that ‘John’ should self-isolate and quality of evidence perception (r=-0.19, *t*=-5.86, df=870, p<.001) as well as perceived trustworthiness (r=-0.27, *t*=-8.19, df=870, p<.001).^1^

### How easy or difficult did participants find the information to understand?

As might be expected, ratings of understanding of the different wordings were associated with the length of the text, ranging from the New Zealand wording being on average rated the hardest to understand, to the US wording on average the easiest.

A two-way ANOVA (test result + test wording) of ratings of ‘how easy’ people found the wording to understand and ‘how completely’ they felt they understood it revealed a main effect of wording (*F*(3,1372) = 7.89, *p* <.001, *η*^*2*^ = 0.017), with post-hoc Tukey tests indicating the New Zealand text was significantly harder to understand than all three other wordings, UK (*p*_adj_=.012), US (*p*_adj_<.001), and experimental wording (*p*_adj_=.009).

An ANOVA of rating of ‘how much effort’ people had to put into understanding the wording revealed a main effect of wording (*F*(3,1381) = 8.53, *p* <.001, *η*^*2*^ = 0.018), with the US wording being significantly lower on invested effort compared to all other groups, UK (*p*_adj_=.045), New Zealand (*p*_adj_<.001), and experimental wording (*p*_adj_=.014), in line with our pilot findings.

## DISCUSSION

Test results are often misinterpreted, particularly if the prevalence of the condition being tested for is very low, where the number of true positives out of the total number of positive tests will be low(13). The difficulty of explaining the effects of prevalence, sensitivity and specificity of a test on the interpretation of a test result has possibly led to caution in what uncertainty is communicated. Added to these complications are the difficulties associated with assessing sensitivity and specificity of COVID-19 swab tests in a real-world context as opposed to a laboratory context, where additional uncertainties are introduced by users carrying out the swabbing themselves.

As a result, the UK’s National Health Service has decided to communicate the results of COVID-19 swab tests with no uncertainty, in an apparent attempt to maximise clarity for the recipient. New Zealand’s Ministry of Health has, by contrast, dedicated an entire website to trying to communicate the uncertainties. The US’s Centers of Disease Control have taken the middle ground, merely adding the caveat that a recipient of a negative test result ‘probably’ doesn’t have COVID-19. These three pieces of information are not comparable in their aims and audience – they are not all information provided to test recipients – but we here compare their effects on the public’s interpretation of test results as a way of gauging how such information affects people’s natural understanding of such a test result.

This study shows that with no additional information about a test result (control group), the UK public demonstrate conservative estimates of the reliability of the results, and have a lower confidence in a negative test result than a positive test result (which is reasonable given that specificity is larger than sensitivity for swab tests, which also holds for ‘quick’ lateral flow tests). The public’s estimate of the reliability and trustworthiness of the results correlates with their reaction to behavioural advice, where again they tend to show a precautionary approach to a negative result, although this may have been because the test recipient had been described as symptomatic and hence ideally the majority of participants would have felt he should self-isolate. The free text that participants gave to explain why they thought the test recipient should or should not self-isolate show how a large number chose to be ‘better safe than sorry’ (although fewer amongst those who were given information about test interpretation from the UK’s NHS website).

The UK’s official, unambiguous, interpretation of the result appeared to lead some participants to interpret a positive result as meaning that the recipient was 100% likely to have COVID-19. That, however, may not be far off the true rate given the low false positive rate for the tests. More worryingly, as some commentators had feared(7), the wording also led some participants to believe that a negative result meant the recipient had a 0% chance of having COVID-19. When it comes to the all-important behavioural interpretation of the results, compared to the control, only the New Zealand wording resulted in a lower proportion of individuals who thought that a (symptomatic) recipient of a negative test result definitely did not need to self-isolate. The experimental information, based on the UK wording but incorporating what we hoped was clearer advice around a negative test result, did not appear to discourage this view. Although the free text responses in this group did seem to suggest an increase in the number of people who picked up on the fact that a symptomatic person should continue to self-isolate.

Both the pilot and the main study found that the test recipient’s chance of having COVID-19, perceived trustworthiness of the result, and perceived quality of evidence of the result were higher for participants viewing positive test results than negative test results. However, the interactions between test result and message wording observed in the pilot did not replicate, and the pilot did not investigate what is arguably the most important set of results: how did participants thought the test recipient should behave after receiving either the positive or negative result. Some of the differences may be due to improvements we made to the questionnaire after the pilot: the difference in how participants were asked to input percentages when asked the test recipient’s chance of having COVID-19 (a slider vs. a text box in the pilot), which made a large difference to the number of participants who provided answers of 50%, and putting the test result in bold to ensure that it was not missed. Nevertheless, in both the pilot and in the main experiment, we found that participants who viewed the New Zealand wording were least likely to provide answers of “50%” when asked the test recipient’s chance of having COVID-19, and were the least likely overall to provide categorical (0% or 100%) responses, suggesting that participants in this wording condition tended to come away with a more nuanced view of what a COVID-19 test result means.

This study has a number of limitations. The scenario given to participants, of a high suspicion (50% chance) that the test recipient had COVID was not clinically likely unless they were a health worker (or already hospitalised because of symptoms) given the prevalence of COVID at the time our study was conducted. We did not introduce either of these factors as we felt they would be likely to bias participants’ interpretation of either the test result or their behavioural recommendation. However, depending on overall prevalence, in many cases the prior probability will be much lower than 50% and we have not tested how this would affect participants’ interpretations. We also presented participants with text taken from official websites, rather than the much shorter text they are likely to receive (e.g. by SMS message) alongside their test result. Additionally, the question asking participants to give a numerical probability of the chances of the likelihood of COVID still resulted in a high proportion of people indicating that they were uncertain how to respond.

Our conclusions, however, are that although the public have a natural sense of the comparative reliability of positive and negative test result, presenting them with an unambiguous interpretation encourages a belief that the results are 100% certain. Advice that a symptomatic person should continue to self-isolate after a single negative test result needs to be given very strongly, in line with UK NHS and government guidance which states that individuals who feel unwell should continue to self-isolate (3,14). We suggest that the UK NHS consider modifying their website text in a similar way to the experimental wording that we tested as this discourages the belief that results are 100% certain. However, it did not appear to affect the belief that a symptomatic person with a negative test result could stop self-isolating. Only the New Zealand wording had this effect, and other agencies might consider adopting wording more similar to that used by the New Zealand Ministry of Health.

## Data Availability

All data and code is available at: https://osf.io/pvhba/

## DATA SHARING

All data and code is available at: https://osf.io/pvhba/.

## ACKNOWLEDGEMENTS AND FUNDING SOURCE

We would like to thank all our participants for giving their time so generously to this project, and to the administrative team who helped run the study.

Funding was provided by the Winton Centre for Risk & Evidence Communication, which is funded from a donation from the David & Claudia Harding Foundation. The Foundation played no role in the study. The authors have no conflicts of interest to declare.

## Supplementary information

### Main variables measured: Main study

| Question | Response options |
| --- | --- |
| How easy or difficult did you find the information above to understand?<br>How completely do you feel you understood the information above?<br>How much effort did you feel you had to put into understanding the information above? | 7-point Likert scale with the ends labelled “None” and “A lot” |
| What is your best guess as to the percent chance that John actually had COVID-19 at the time of his test, given his test result? | Slider scale with 10% markings with the ends labelled “0% = John definitely **didn't** have COVID-19” and “100% = John definitely **did** have COVID-19”<br>Recorded as an integer between 0-100 |
| How sure or unsure are you about your answer above, regarding the percent chance that John actually had COVID-19? | Slider scale with the ends labelled “very unsure” and “very sure”<br>Recorded as an integer between 0-100 |
| How high or low do you think the quality of the evidence behind John's test result is? | Slider scale with the ends labelled “very low” and “very high”<br>Recorded as an integer between 0-100 |
| To what extent do you think that the result from John’s test was certain or uncertain? | Slider scale with the ends labelled “very uncertain” and “very certain”<br>Recorded as an integer between 0-100 |
| To what extent do you think that John's test result was...<br>Accurate?<br>Reliable?<br>Trustworthy? | 7-point Likert scale with the ends labelled "Not at all" and "Very much" |
| Given John's test result, how much do you agree or disagree with the statement that "John should now isolate himself from other people"? | Slider scale with the ends labelled "completely disagree" and "completely agree"<br>Recorded as an integer between 0-100 |
| What are your reasons for your answer above? | Free text |
| To what extent do you think the people who are responsible for communicating the results of the COVID-19 swab tests are trustworthy? | 7-point Likert scale with the ends labelled "Not at all" and "Very much" |
| To what extent do you think policies and restrictions, such as rules for being allowed to enter a country, should be based on the results of a swab test? | 7-point Likert scale with the ends labelled "Not at all" and "Very much" |

### Stimulus 1 (United States CDC)

#### COVID-19 Testing Overview

Two kinds of tests are available for COVID-19: viral tests and antibody tests.

- A viral test tells you if you have a current infection.
- An antibody test might tell you if you had a past infection.

#### Considerations for who should get tested

- People who have symptoms of COVID-19
- People who have had close contact (within 6 feet of an infected person for at least 15 minutes) with someone with confirmed COVID-19
- People who have been asked or referred to get testing by their healthcare provider, local icon or state health department

Not everyone needs to be tested. If you do get tested, you should self-quarantine/isolate at home pending test results and follow the advice of your health care provider or a public health professional.

#### How to get tested for current COVID-19 infection

- You can visit your state or local icon health department’s website to look for the latest local information on testing.
- If you have symptoms of COVID-19 and want to get tested, call your healthcare provider first.

#### Results

- If you test positive, know what protective steps to take to prevent others from getting sick.
- If you test negative, you probably were not infected at the time your sample was collected. The test result only means that you did not have COVID-19 at the time of testing. Continue to take steps to protect yourself.

### Stimulus 2 (United Kingdom NHS)

#### Your coronavirus test result

This information is about the swab test to check if you have coronavirus (COVID-19).

#### Getting your test result

You’ll get a text or email when your result is ready.

Most people get their test results the day after taking the test. Some results might take longer, but you should get them in 72 hours.

There are 3 types of result you can get:

- negative
- positive
- unclear, void, borderline or inconclusive

#### Stay at home

If you had a test because you had symptoms, you and anyone you live with must stay at home (self-isolate) until you get your result.

Anyone in your support bubble must also self-isolate until you get your result.

#### Negative test result

A negative result means the test did not find coronavirus.

You do not need to self-isolate if your test is negative, as long as:

- everyone you live with who has symptoms tests negative
- everyone in your support bubble who has symptoms tests negative
- you were not told to self-isolate for 14 days
- you feel well – if you feel unwell, stay at home until you’re feeling better
- If you have diarrhoea or you’re being sick, stay at home until 48 hours after they’ve stopped.

If you’re a health or care worker, check with your employer before going back to work.

#### Positive test result

A positive result means you had coronavirus when the test was done. If your test is positive, you must self-isolate immediately.

- If you had a test because you had symptoms, keep self-isolating for at least 10 days from when your symptoms started.
- If you had a test but have not had symptoms, self-isolate for 10 days from when you had the test.

Anyone you live with, and anyone in your support bubble, must self-isolate for 14 days from when you start self-isolating.

### Unclear, void, borderline or inconclusive test result

An unclear, void, borderline or inconclusive result means it’s not possible to say if you had coronavirus when the test was done.

Get another coronavirus test as soon as possible if this happens.

If you had a test because you had symptoms, you must keep self-isolating and have another test within 5 days of your symptoms starting.

If you’re not able to have another test in time, you must self-isolate for at least 10 days from when your symptoms started. Anyone you live with, and anyone in your support bubble, must self-isolate for 14 days.

If you had a test but have not had any symptoms, you do not need to self-isolate while you wait to get another test. People you live with, and anyone in your support bubble, do not need to self-isolate.

### Stimulus 3 (New Zealand Ministry of Health)

#### COVID-19 test results and their accuracy

Information about how COVID-19 test results are reported, why they are sometimes uncertain, and what they can and cannot tell us

#### How the results are reported

COVID-19 test results are reported as positive or negative. If the test result is:

- positive - the virus (its genes) was detected in the sample
- negative - the virus was not detected in the sample.

Sometimes the test result might be reported as a ‘weak positive’, which means a very small amount of virus is in the sample. This could mean the person was tested towards the end of the illness when the level of virus in their body was low.

#### The accuracy of test results

The viral test for COVID-19 is accurate when taken in ideal conditions. A recent laboratory study found that different COVID-19 testing kits correctly detected COVID-19 in samples more than 95% (and frequently 100%) of the time. When tests were done on samples without the virus, the tests correctly gave a negative result 96% of the time.

But it is important to remember that tests don’t work as well in the real world. No viral test is 100% accurate. In real-world use, the viral test for COVID-19 is not 100% ‘sensitive’ (meaning able to correctly identify people with the disease all of the time). This means that if 100 people who have the disease are tested – some will have a negative result (i.e. a false negative result).

Reasons for a false negative test result could be because:

- the sample was taken at the wrong time (too early or too late)
- the swab did not pick up any pieces of the virus, or
- the sample of mucus (or liquid from the lungs) wasn’t big enough.

This means that it is very important to isolate yourself if a health professional asks you to, even if your test result is negative.

The viral test for COVID-19 is much better at correctly identifying people who don’t have COVID-19 (this is known as a higher ‘specificity’). We expect very few (if any) false positive test results (a false positive being a positive test result for someone who does not have the disease).

#### What the test results can and cannot tell us

Even when we take the uncertainties of testing into account, the results can tell us a few things.

A positive test tells us that a person either has COVID-19 (whether they have symptoms, or not) or has had COVID-19 recently. We may not be able to distinguish whether the person is currently infectious or not so we will take a precautionary approach.

A positive test cannot tell us:

- if the person is currently infectious
- how ill the person is likely to become.

A negative test can tell us:

- the person was unlikely to be infectious at the time of the test.

A negative test cannot tell us:

- if the person was exposed to the virus or not
- if they are in the early stages of incubating the disease
- if they caught COVID-19 in the past
- if they were infectious in the past
- that they will not get COVID-19 in the future.

Note: if a person has a negative test result and they are at a higher risk of having COVID-19, they may be tested again.

### Stimulus 4 (‘experimental’ based on UK NHS) – Main study only

#### Your coronavirus test result

This information is about the swab test to check if you have coronavirus (COVID-19).

#### Getting your test result

You’ll get a text or email when your result is ready.

Most people get their test results the day after taking the test. Some results might take longer, but you should get them in 72 hours.

There are 3 types of result you can get:

- negative - the virus was not detected in the sample.
- positive - the virus was detected in the sample.
- unclear, void, borderline or inconclusive - it was not possible to provide a result for your sample.

Based on data about the test procedure we use, we expect very few (if any) people being given a ‘positive’ test result if they do not have the disease.

Whilst data shows the test is even better at correctly identifying people who don’t have COVID-19, there are other reasons why some people may be given a ‘negative’ result when they do actually have COVID-19, such as:

- the sample was taken at the wrong time (too early or too late)
- the swab did not pick up any pieces of the virus, or
- the sample of mucus (or liquid from the lungs) wasn’t big enough.

This means that it is very important to follow the instructions below, even if your test result is negative.

#### Stay at home

If you had a test because you had symptoms, you and anyone you live with must stay at home (self-isolate) until you get your result.

Anyone in your support bubble must also self-isolate until you get your result.

#### Negative test result

A negative result means the test did not find coronavirus.

You do not need to self-isolate if your test is negative, unless:

- you have any symptoms – if you feel unwell, stay at home until you’re feeling better
- someone you live with, or who is in your support bubble, has symptoms and has not yet been tested
- someone you live with, or who is in your support bubble, has tested positive
- you have been told to self-isolate for 14 days
- you have diarrhoea or you’re being sick. If this is the case, stay at home until 48 hours after they’ve stopped.

If you’re a health or care worker, check with your employer before going back to work.

#### Positive test result

A positive result means you almost certainly had coronavirus when the test was done.

If your test is positive, you must self-isolate immediately.

- If you had a test because you had symptoms, keep self-isolating for at least 10 days from when your symptoms started.
- If you had a test but have not had symptoms, self-isolate for 10 days from when you had the test.

Anyone you live with, and anyone in your support bubble, must self-isolate for 14 days from when you start self-isolating.

#### Unclear, void, borderline or inconclusive test result

An unclear, void, borderline or inconclusive result means it’s not possible to say if you had coronavirus when the test was done.

Get another coronavirus test as soon as possible if this happens.

If you had a test because you had symptoms, you must keep self-isolating and have another test within 5 days of your symptoms starting.

If you’re not able to have another test in time, you must self-isolate for at least 10 days from when your symptoms started. Anyone you live with, and anyone in your support bubble, must self-isolate for 14 days.

If you had a test but have not had any symptoms, you do not need to self-isolate while you wait to get another test. People you live with, and anyone in your support bubble, do not need to self-isolate.

## Pilot study

### Hypotheses

The first hypothesis preregistered in the pilot study (H1) was that real-world messaging which clearly acknowledges the presence of uncertainty (Ministry of Health New Zealand advice) would facilitate a better understanding of the true level of uncertainty associated with positive and negative PCR test results than messaging that does not (UK National Health Service advice).

The second preregistered hypothesis (H2) was that reading messaging which defines the meaning of positive and negative test results but fails to acknowledge the presence of uncertainty (UK National Health Service advice) would lead participants to underestimate the true probability of having COVID-19 after a negative test result. The full preregistration can be found at https://osf.io/8n62f.

### Methods

#### Procedure (preregistered)

As in the main study, participants were recruited from two sources, Prolific and Respondi, and presented with a questionnaire in Qualtrics. They were randomised to their condition via the Qualtrics ‘randomise’ function. After consenting to take part, they were presented with a scenario in which a fictional individual (‘John’) has been feeling ill. Participants were told that based on symptoms alone, a knowledgeable doctor believes that this fictional individual has a 50-50 chance of having COVID-19. They were then told the result of this individual’s COVID-19 viral test result (either positive or negative). Depending on condition, they were then also presented with information about interpreting a COVID-19 test result published by the U.S. Centers for Disease Control, the U.K. National Health Service, the New Zealand Ministry of Health, or no information at all (see above for full text of each). That is, each participant was randomly assigned to one of eight conditions in a between-subjects 2 (test result: positive, negative) x 4 (message: CDC, NHS, Ministry of Health NZ, or none) factorial design. Participants who failed an attention check question were excluded. After reading the information, participants were asked the following questions:

| Question | Response options |
| --- | --- |
| How easy or difficult did you find the information above to understand? | 7-point Likert scale with the ends labelled "None" and "A lot" |
| How completely do you feel you understood the information above? |  |
| How much effort did you feel you had to put into understanding the information above? |  |
| What is your best guess as to the percent chance that John actually had COVID-19 at the time of his test, given his test result?<br>(Please type a number without the '%' sign) | Free-text (limited to numerical answers only) |
| How high or low do you think the quality of the evidence for the accuracy of 'PCR' or 'swab' testing in general is? | Slider scale with the ends labelled "very low" and "very high"<br>Recorded as an integer between 0-100 |
| How high or low do you think the quality of the evidence behind John's test result is? | Slider scale with the ends labelled "very low" and "very high"<br>Recorded as an integer between 0-100 |
| To what extent do you think that the result from John's test was certain or uncertain? | Slider scale with the ends labelled "very uncertain" and "very certain"<br>Recorded as an integer between 0-100 |
| To what extent do you think that John's test result was...<br>Accurate?<br>Reliable?<br>Trustworthy? | 7-point Likert scale with the ends labelled "Not at all" and "Very much" |
| To what extent do you think the people who are responsible for communicating the results of the COVID-19 'PCR' or 'swab' tests are trustworthy? | 7-point Likert scale with the ends labelled "Not at all" and "Very much" |
| To what extent do you think policies and restrictions, such as rules for being allowed to enter a country, should be based on the results of a 19 'PCR' or 'swab' test? | 7-point Likert scale with the ends labelled "Not at all" and "Very much" |

Prior research (van der Bles et al., 2020) suggested that explicitly communicating uncertainty increases the degree to which participants perceive a number as uncertain, with effect sizes from *d* =.37 to.72, although we expected we may see smaller effect sizes given differences in our experimental task and setup. Our power analysis was based on the tests for which we have clear hypotheses. For Analysis 1, to achieve 95% power with an alpha of.05, d =.3 for those tests, power analysis in G*Power indicated that we would require 1176 participants. To account for attrition due to failing of the attention check and to buffer for potentially smaller observed effect sizes of the effects for which we do not have clear directional hypotheses or prior research findings, we sampled a total of 1850 participants.

### Analyses (preregistered)

#### Analysis 1a

Analysis 1a, the preregistered analysis intended to test H1, was a 2 (test result) x 4 (message) factorial ANOVA, with participants’ understanding of the predictive value of the test result as the dependent variable, with a planned comparison comparing participants’ understanding of the predictive value of the test result for participants viewing the NHS message vs. the Ministry of Health NZ message.

The key dependent variable for this analysis, “*participants’ understanding of the predictive value of the test result*,” was operationalized as the difference between each participant’s estimate of the probability of COVID (“What is your best guess as to the percent chance that John actually had COVID-19 at the time of his test?”) and an expert estimate, i.e., the estimate of the posterior probability of having COVID-19 (corresponding to a prior of 50%) as determined by Watson, Whiting & Brush (2020), Table 1: 93% for a positive test result, and 24% for a negative test result.

#### Analysis 1b

The preregistered analysis corresponding to H2 was a one-sample t-test comparing participants’ estimates to the expert estimates, for participants who saw the negative test result and the NHS message. While this hypothesis was restricted to participants in the negative test result condition who viewed the NHS message, we noted in the preregistration that we would conduct similar tests for participants who saw the negative test result as well as either the CDC message, Ministry of Health NZ message, or no message; we had no clear hypothesis about whether participants would underestimate the probability of COVID in these conditions.

#### Analysis 2

We preregistered 2 (test result) x 4 (message) factorial ANOVAs with participants’ understanding, quality of evidence perception, perceived trustworthiness, and decision making as the dependent variables. We identified a number of measures that we intended to use to operationalize these constructs. We noted in our preregistration that if correlations between measures corresponding to the same construct were at least 0.7 and Cronbach’s alpha was sufficient, we would combine multiple items into a single index for that construct. The measures are shown in the table above.

### Results

Due to slight accidental oversampling 1869 participants were sampled, of whom 1657 passed the attention check and were included in analyses. 895 were recruited via Prolific and 762 from Respondi.

#### Analysis 1a

The ANOVA found a main effect of test result, F(1, 1649) = 137.6, *p* <.001, *η*_*p*_^*2*^ =.077, with participant estimates being farther from the expert estimate for positive test results (M = 33.7%, SD = 20.8%) than for negative test results (M = 24.1%, SD = 12.3%). This may have been influenced by the fact that half (50.0%) of participants provided an answer of 50% as their estimate—a response that other research has found participants commonly use to indicate “I don’t know” rather than to indicate their true best guess (De Bruin et al., 2000)— and 50% was closer to the expert estimate for negative test results. There was also a main effect of message, F(3, 1649) = 12.1, *p* <.001, *η*_*p*_^*2*^ =.022; differences between participant estimates and expert estimates are given in Table S1. The planned comparison comparing participants’ understanding of the predictive value of the test result for participants viewing the UK National Health Service message vs. the Ministry of Health NZ message found a greater discrepancy between participant and expert estimates for those viewing the UK message (M = 31.7%, SD = 16.4%) vs. those viewing the NZ message (M = 27.2%, SD = 19.3%), *t*(1649) = 3.93, *p* <.001.

In addition to this difference between those viewing the NZ and UK messages, Tukey’s post-hocs showed that differences between UK/US, UK/control, US/control, and US/NZ were all significant, *p*_*adj*_ <.05. There was also a significant interaction, F(3, 1649) = 19.8, p <.001, *η* _*p*_^*2*^=.035, Table S1. For example, for those who were told John’s test result was positive, the difference between expert and participant estimates was significantly greater for participants who had seen the US wording than for the control group (*p*_*adj*_ <.001), but there was no such difference for those who were told John’s test result was negative (*p*_*adj*_ *=* 1.0).

**Table S1.** Differences between participant and expert estimates by wording group and test result.

| *Message* | *Difference between participant and expert estimates, Mean (SD)* |  |  |
| --- | --- | --- | --- |
| UK | 31.7% (16.4%) | *Negative test results* | 23.7% (10.6%) |
|  |  | *Positive test results* | 39.7% (17.3%) |
| US | 30.8% (18.2%) | *Negative test results* | 23.2% (9.1%) |
|  |  | *Positive test results* | 38.3% (21.6%) |
| NZ | 27.2% (19.3%) | *Negative test results* | 26.8% (16.0%) |
|  |  | *Positive test results* | 27.5% (22.4%) |
| Control | 25.9% (16.2%) | *Negative test results* | 22.4% (11.7%) |
|  |  | *Positive test results* | 29.0% (19.0%) |

Given that the distribution was highly non-normal, due to a high proportion of participants estimating John’s chance of COVID to be 50%, we also conducted a non-parametric aligned ranks transformation ANOVA, which also found both main effects and the interaction to be significant.

#### Analysis 1b

For an individual having a 50% prior probability of COVID, who then tested negative, the expert estimate of that individual’s probability of having COVID despite the negative test is 24%. Participants who were told that John had tested negative had significantly higher mean estimates than this irrespective of message condition (UK message: 39.4%, *p* <.001; US message: 36.7%, *p* <.001; NZ message: 41.5%, *p* <.001; control: 38.3%, *p* <.001). Participants who were told that John had tested *positive* had *lower* mean estimates than the expert estimate of 93% (UK message: 54.1%, *p* <.001; US message: 56.3%, *p* <.001; NZ message: 67.3%, *p* <.001; control: 65.9%, *p* <.001).

#### Analysis 2

Answers to the questions ‘How easy or difficult did you find the information that we just showed you to understand?’ and ‘How completely do you feel you understood the information?’ were highly correlated (r =.81) and combined into an index. The effort investment item, “How much effort did you feel you had to put into understanding the information above?”, was correlated at less than.40 for both ease and completeness of understanding, and analysed as a single item. The ANOVA on the ease and completeness of understanding index measure revealed a main effect of message, F(2, 1236) = 18.6, p <.001, *η* _*p*_^*2*^ =.029. There was no main effect of test result and no interaction. Post-hoc Tukey tests suggested that participants felt the NZ message (M = 5.03, SD = 1.38) and the UK message (M = 5.19, SD = 1.49) were more difficult to understand than the US message (M = 5.61, SD = 1.31), both p <.001. The ANOVA on the effort invested for understanding measure likewise revealed a main effect of message, F(2, 1238) = 14.88, p <.001, *η*_*p*_^*2*^ =.023. There was no main effect of test result and no interaction. Post-hoc Tukey tests suggested that participants felt they had to invest more effort into understanding the NZ message (M = 4.91, SD = 1.58) and the UK message (M = 4.69, SD = 1.86) compared to the US message (M= 4.25, SD = 1.85), both p <.001. The control condition was not included in these analyses as participants were not asked these questions in the control condition.

With respect to perceptions of the quality of the evidence for the accuracy of ‘PCR’ or ‘swab’ testing generally *(‘How high or low do you think the quality of the evidence for the accuracy of ‘PCR’ or ‘swab’ testing in general is?’)*, there was a main effect of test result, F(1, 1648) = 19.0, p <.001, *η*_*p*_^*2*^ =.011, with higher quality perceived by those who saw a positive test result (M = 73.0, SD = 19.3) vs. those shown a negative test result (M = 68.7, SD = 20.9). There was also an interaction between test result and message, F(3, 1648) = 2.6, p =.049, *η*_*p*_^*2*^ =.005; post-hoc Tukey tests suggested those shown the positive result perceived higher quality than those shown the negative result in the control condition (p <.001) and in the NZ condition (p =.03), but the differences were not significant in the UK and US conditions. There was no main effect of message.

With respect to the specific test result they were provided *(‘How high or low do you think the quality of the evidence behind John’s test result is?’)*, there was a main effect of test result, F(1, 1646) = 33.5, p <.001, *η*_*p*_^*2*^ =.020, with higher quality perceived by those who saw a positive test result (M = 73.3, SD = 18.7) vs. those shown a negative test result (M = 67.8, SD = 20.5). There was also an interaction between test result and message, F(3, 1646) = 5.2, p =.002, *η*_*p*_^*2*^ =.009. Post-hoc Tukey tests comparing the dependent variable for the different levels of test result, conditioned on message, suggested that positive tests were perceived as higher-quality than negative tests in the control condition (p <.001) and in the NZ condition (p =.003), but the differences were not significant in the UK and US conditions. There was no main effect of message.

Answers to the question ‘*To what extent do you think that the result from John’s test was certain or uncertain?*’ and to the questions ‘*To what extent do you think that John’s test result was trustworthy?’/’accurate?’/’reliable?*’ were all highly correlated (all r > 0.7, Cronbach’s alpha = 0.95); these were therefore rescaled to 1-7 scales and averaged into a single “perceived trustworthiness” index. Answers to the question ‘*To what extent do you think the people who are responsible for communicating the results of the COVID-19 ‘PCR’ or ‘swab’ tests are trustworthy?*’ were much less correlated and this question was therefore not included in the index. The perceived trustworthiness ANOVA revealed a main effect of test result, F(1, 1643) = 47.3, p <.001, *η*_*p*_^*2*^ =.028, with higher trustworthiness for those who saw a positive test result (M = 5.37, SD = 1.14) vs. those shown a negative test result (M = 4.96, SD = 1.27). There was also an interaction between test result and message, F(3, 1643) = 4.0, p =.007, *η*_*p*_^*2*^ =.007. Post-hoc Tukey tests comparing the dependent variable for the different levels of test result, conditioned on message, suggested that those shown the positive result reported higher trust than those shown the negative result in the control (p <.001), NZ (p <.001), and US (p =.005) conditions, but the difference was not significant in the UK condition. There was no main effect of message.

Finally, concerning responses to the question *‘To what extent do you think policies and restrictions, such as rules for being allowed to enter a country, should be based on the results of a ‘PCR’ or ‘swab’ test?’*, there was a main effect of test result, F(1, 1652) = 6.5, p =.01, *η*_*p*_^*2*^ =.004, with greater agreement for those told that John’s test result was positive (M = 5.42, SD = 1.45) than for those told it was negative (M = 5.23, SD = 1.48). There was also a main effect of message, F(3, 1652) = 2.9, p =.04, *η*_*p*_^*2*^ =.005. There was no interaction, and differences between message conditions (NZ mean 5.21, SD 1.53; control mean 5.23, SD 1.46; UK mean 5.42, SD 1.44; US mean 5.44, SD 1.43) were not significant in a post-hoc Tukey test that included all six pairwise comparisons.

### Non-preregistered analyses

#### Estimates of John’s true probability of having COVID-19

While Analysis 1a considered the absolute difference between participant and expert estimates, participants’ actual estimates of John’s probability of having COVID-19 are also of interest. While we had investigated these to some degree in Analysis 1b, we felt it worthwhile to conduct an exploratory 2×4 ANOVA to look for potential differences by test result and by message. Nonparametric aligned ranks transformation ANOVA was chosen due to the non-normality of the distribution. There was a main effect of test result, F(1, 1649) = 392.6, p <.001, *η* _*p*_^*2*^ =.925, with participants estimating John to have a higher probability of having COVID-19 if his test result was positive (mean = 60.9%, SD = 23.1%) than if it was negative (mean = 39.0%, SD = 22.5%). There was also a main effect of wording, F(3, 1649) = 9.9, *η*_*p*_^*2*^ =.535, and a significant interaction, F(3, 1649) = 8.6, p <.001. Nonparametric post-hoc tests (Mann-Whitney U tests) suggested that averaged across test result, differences between all pairs of wording groups were significant (all *p* <.001, Table S2), with the exception of the UK/US and control/NZ comparisons.

Zooming in on participants who were told that John’s test result was positive, nonparametric post-hocs suggested differences between NZ/US, NZ/UK, control/US, and control/UK (all p <.001), with estimates for the NZ and control groups higher than those for the US and the UK (Table S2). Differences among answers for those told that John’s test result was negative were not nearly so stark, with unadjusted nonparametric post-hocs only finding a possible difference between the NZ and US estimates (Table S2), p =.04.

**Table S2.** Participant estimates of the probability that John has COVID-19, by wording group and test result.

| Message | Participant estimates of the probability that John has COVID-19, Mean (SD) |  |  |
| --- | --- | --- | --- |
| UK | 46.8% (21.4%) | Negative test result | 39.5% (20.9%) |
|  |  | Positive test result | 54.2% (19.2%) |
| US | 46.6% (24.9%) | Negative test result | 36.7% (21.5%) |
|  |  | Positive test result | 56.3% (24.1%) |
| NZ | 53.8% (28.3%) | Negative test result | 41.5% (25.9%) |
|  |  | Positive test result | 67.3% (24.5%) |
| Control | 52.7% (25.4%) | Negative test result | 38.3% (20.9%) |
|  |  | Positive test result | 65.9% (21.7%) |

Further exploratory analysis revealed that the distributions behind the means of each wording group were different. Overall, participants differed in their propensity to give a categorical ‘100%’ or ‘0%’ answer by message condition; a chi-squared test comparing participants giving categorical vs. noncategorical answers by wording condition was significant, *X*^2^(3, *N* = 1657) = 16.8, with the NZ wording group having the lowest propensity to answer categorically overall, although results were not consistent when split out into participants in the positive vs. negative test result conditions.

#### Who was most likely to answer 50%?

As mentioned, half (50.0%) of participants provided an answer of 50% as their estimate of John’s probability of having COVID-19. These participants may simply have been indicating that they did not know the answer (De Bruin et al., 2000), or they may have felt that the test result was completely uninformative (or simply been confused by it) and therefore reproduced the pre-test percentage. In any of these cases, answering precisely 50% seems to be indicative of a lack of understanding that John’s probability of having COVID-19 should be adjusted upwards on receipt of a positive test result and downwards on a negative result. We therefore felt it worthwhile to look for differences across message conditions with respect to participants’ propensity to answer “50%,” and indeed a *X*^*2*^ test showed that the number of participants answering 50% vs. any other value differed by condition, (*X*^2^(3, *N* = 1657) = 48.1, *p* <.001, Table S3). Follow-up *X*^*2*^ tests comparing the UK message condition to each other group in turn found that those in the UK message group were more likely to answer 50% than those in the US (*p* =.02), NZ (*p* <.001), or Control (*p* <.001) message groups.

**Table S3.** Number of participants answering 50% when asked to express the probability that John has COVID-19, by wording group and test result.

| *Message* | *Number of participants answering 50% (percentage)* |  |  |
| --- | --- | --- | --- |
| UK | 260/420 (61.9%) | *Negative test results* | 119/210 (56.7%) |
|  |  | *Positive test results* | 141/210 (67.1%) |
| US | 220/411 (53.5%) | *Negative test results* | 105/204 (51.5%) |
|  |  | *Positive test results* | 115/207 (55.6%) |
| NZ | 164/412 (39.8%) | *Negative test results* | 90/215 (41.9%) |
|  |  | *Positive test results* | 74/197 (37.5%) |
| Control | 184/414 (44.4%) | *Negative test results* | 88/198 (44.4%) |
|  |  | *Positive test results* | 96/216 (44.4%) |

## CONSORT 2010 checklist of information to include when reporting a randomised trial*

CONSORT 2010 checklist of information to include when reporting a randomised trial\*
| Section/Topic | Item No | Checklist item | Reported on page No |
| --- | --- | --- | --- |
| **Title and abstract** |  |  |  |
|  | 1a | Identification as a randomised trial in the title | 1 |
|  | 1b | Structured summary of trial design, methods, results, and conclusions (for specific guidance see CONSORT for abstracts) | 2-3 |
| **Introduction** |  |  |  |
| Background and objectives | 2a | Scientific background and explanation of rationale | 4-6 |
|  | 2b | Specific objectives or hypotheses | 6, 11-12 |
| **Methods** |  |  |  |
| Trial design | 3a | Description of trial design (such as parallel, factorial) including allocation ratio | 7-10 |
|  | 3b | Important changes to methods after trial commencement (such as eligibility criteria), with reasons | NA |
| Participants | 4a | Eligibility criteria for participants | 7 |
|  | 4b | Settings and locations where the data were collected | 7 |
| Interventions | 5 | The interventions for each group with sufficient details to allow replication, including how and when they were actually administered | 7-10 & supplementary info |
| Outcomes | 6a | Completely defined pre-specified primary and secondary outcome measures, including how and when they were assessed | 7-10 & supplementary info |
|  | 6b | Any changes to trial outcomes after the trial commenced, with reasons | NA |
| Sample size | 7a | How sample size was determined | 10 |
|  | 7b | When applicable, explanation of any interim analyses and stopping guidelines | NA |
| **Randomisation:** |  |  |  |
| Sequence generation | 8a | Method used to generate the random allocation sequence | 7 |
|  | 8b | Type of randomisation; details of any restriction (such as blocking and block size) | NA |
| Allocation concealment mechanism | 9 | Mechanism used to implement the random allocation sequence (such as sequentially numbered containers), describing any steps taken to conceal the sequence until interventions were assigned | NA |
| Implementation | 10 | Who generated the random allocation sequence, who enrolled participants, and who assigned participants to interventions | 7 |
| Blinding | 11a | If done, who was blinded after assignment to interventions (for example, participants, care providers, those assessing outcomes) and how | NA |
|  | 11b | If relevant, description of the similarity of interventions | NA |
| Statistical methods | 12a | Statistical methods used to compare groups for primary and secondary outcomes | 13-21 |
|  | 12b | Methods for additional analyses, such as subgroup analyses and adjusted analyses | 13-21 |
| **Results** |  |  |  |
| Participant flow (a diagram is strongly recommended) | 13a | For each group, the numbers of participants who were randomly assigned, received intended treatment, and were analysed for the primary outcome | 10 |
|  | 13b | For each group, losses and exclusions after randomisation, together with reasons | 10 |
| Recruitment | 14a | Dates defining the periods of recruitment and follow-up | NA |
|  | 14b | Why the trial ended or was stopped | NA |
| Baseline data | 15 | A table showing baseline demographic and clinical characteristics for each group | Table 1 (p7-8) |
| Numbers analysed | 16 | For each group, number of participants (denominator) included in each analysis and whether the analysis was by original assigned groups | 10 |
| Outcomes and estimation | 17a | For each primary and secondary outcome, results for each group, and the estimated effect size and its precision (such as 95% confidence interval) | 13-21 |
|  | 17b | For binary outcomes, presentation of both absolute and relative effect sizes is recommended | NA |
| Ancillary analyses | 18 | Results of any other analyses performed, including subgroup analyses and adjusted analyses, distinguishing pre-specified from exploratory | 14, 16-21, supplementary information |
| Harms | 19 | All important harms or unintended effects in each group (for specific guidance see CONSORT for harms) | NA |
| **Discussion** |  |  |  |
| Limitations | 20 | Trial limitations, addressing sources of potential bias, imprecision, and, if relevant, multiplicity of analyses | 24+25 |
| Generalisability | 21 | Generalisability (external validity, applicability) of the trial findings | 25 |
| Interpretation | 22 | Interpretation consistent with results, balancing benefits and harms, and considering other relevant evidence | 22-25 |
| **Other information** |  |  |  |
| Registration | 23 | Registration number and name of trial registry | 3 |
| Protocol | 24 | Where the full trial protocol can be accessed, if available | 3 |
| Funding | 25 | Sources of funding and other support (such as supply of drugs), role of funders | 26 |

\*We strongly recommend reading this statement in conjunction with the CONSORT 2010 Explanation and Elaboration for important clarifications on all the items. If relevant, we also recommend reading CONSORT extensions for cluster randomised trials, non-inferiority and equivalence trials, non-pharmacological treatments, herbal interventions, and pragmatic trials. Additional extensions are forthcoming: for those and for up to date references relevant to this checklist, see www.consort-statement.org.

## Footnotes

1 Non-parametric correlation analyses were conducted as a robustness check. Results were in line with the parametric analyses.

